# Addition of Bupropion or Varenicline to Nicotine Replacement Therapy After Acute Coronary Syndrome: A Propensity-Matched Real-World Analysis

**DOI:** 10.64898/2026.04.21.26351432

**Authors:** Abdul Qadeer, Najam Gohar, Pankti Maniyar, Lorena Mata Juarez, Nimra Shafi, Shiva Raju Gollapally Krishna, Ibrahim Mortada, Quinn R Pack, Hani Jneid, Diann E. Gaalema

**Affiliations:** Department of Cardiovascular Medicine, University of Texas Medical Branch, Galveston, TX, USA; Department of Internal Medicine, Ameer-ud-Din Medical College, Lahore, Pakistan; Arnot Ogden Medical Center, Elmira, NY, USA; Department of Internal Medicine, University of Texas Medical Branch, Galveston, TX, USA; Sainy Mary Medical Center, PA USA; Department of Healthcare Delivery and Population Sciences, University of Massachusetts Chan Medical School – Baystate, Springfield, MA

**Keywords:** ACS, acute coronary syndrome, smoking-cessation, TriNetX database, propensity score matching

## Abstract

**Introduction:** Smoking cessation after acute coronary syndrome (ACS) is a Class I recommendation, yet prescription pharmacotherapy use remains low and its real-world cardiovascular effectiveness when added to nicotine replacement therapy (NRT) is poorly characterized.

**Methods:** We conducted a retrospective cohort study using the TriNetX US Collaborative Network (67 healthcare organizations). Adults hospitalized with ACS who received NRT within one month, serving as a proxy for active smoking status, were identified. Two co-primary propensity-matched (1:1, 50 covariates, caliper 0.10 SD) comparisons evaluated bupropion + NRT and varenicline + NRT individually versus NRT alone; a supportive analysis evaluated combined pharmacotherapy versus NRT alone. All-cause mortality was the primary endpoint. Secondary outcomes included MACE, heart failure exacerbations, major bleeding, TIA/stroke, emergency rehospitalizations, and cardiac rehabilitation utilization, assessed at 6 months and 1 year via Kaplan-Meier analysis. Hazard ratios (HRs) greater than 1.0 indicate higher hazard in the NRT-only group.

**Results:** After matching, the combined analysis comprised 8,574 pairs, the bupropion analysis 4,654 pairs, and the varenicline analysis 2,126 pairs. At 1 year, the combined pharmacotherapy group had significantly lower all-cause mortality (HR 1.26, 95% CI 1.16-1.37), MACE (HR 1.16, 95% CI 1.12-1.21), heart failure exacerbations (HR 1.16, 95% CI 1.08-1.25), major bleeding (HR 1.18, 95% CI 1.08-1.28), and greater cardiac rehabilitation utilization (HR 0.82, 95% CI 0.74-0.92; all p < 0.001). TIA/stroke did not differ significantly. Six-month results were consistent. Both varenicline and bupropion individually showed lower mortality and MACE. A urinary tract infection falsification endpoint showed no between-group differences, supporting matching validity. The pharmacotherapy group had higher rates of new-onset depression, driven predominantly by bupropion recipients.

**Conclusions:** In this propensity-matched real-world analysis, adding prescription smoking cessation pharmacotherapy to NRT after ACS was associated with lower mortality and fewer adverse cardiovascular events, supporting broader integration into post-ACS care pathways.

## INTRODUCTION

Cigarette smoking remains one of the most significant and modifiable risk factors for cardiovascular morbidity and mortality, particularly among individuals with established coronary artery disease (1,2). Persistent smoking after an acute coronary syndrome (ACS) is associated with a substantially increased risk of recurrent ischemic events, progression of heart failure, bleeding complications, and premature death (3,4). Smoking cessation following ACS represents a cornerstone of secondary prevention and is strongly endorsed in contemporary cardiovascular prevention strategies, offering a distinct opportunity to initiate smoking cessation strategies during hospitalization and improve long-term outcomes (5–7).

Pharmacologic therapies, such as nicotine replacement therapy (NRT), varenicline, and bupropion, are effective tools to facilitate smoking cessation (8). NRT alleviates withdrawal symptoms and cravings, particularly when initiated during or shortly after ACS hospitalization, while bupropion and varenicline act via complementary mechanisms to enhance abstinence (9). Despite strong clinical evidence from clinical trials and real-world studies, these therapies remain substantially underutilised: only ∼20% of post-ACS patients receive NRT, varenicline is prescribed in <1%, and in-hospital bupropion shows limited effectiveness (3,10). Barriers include concerns about cardiovascular safety, clinical inertia, and lack of routine acute-care prescribing, highlighting a persistent gap between guideline recommendations and practice.

Randomized trials indicate that combining NRT with non-nicotine pharmacotherapies improves long-term abstinence compared with NRT alone (11). Varenicline, in particular, has demonstrated high efficacy, and prior concerns regarding cardiovascular and neuropsychiatric risks have largely been mitigated, as shown in the EAGLES trial, leading to the removal of the FDA black box warning (12). Nevertheless, evidence is limited on the real-world safety and effectiveness of adding varenicline or bupropion to NRT in patients hospitalised with ACS. Most data come from controlled trials with selective populations, leaving uncertainty about how these therapies perform across broader, high-risk patient cohorts encountered in routine clinical practice (3).

Addressing this knowledge gap is critical, as understanding whether combination pharmacotherapy confers incremental cardiovascular benefits without increasing adverse events can guide clinical decision-making and improve uptake of evidence-based cessation strategies. Real-world data can complement trial evidence by capturing diverse patient populations, comorbidities, and typical clinical use patterns, offering insights into both effectiveness and safety in routine practice.

In light of these considerations, we conducted a retrospective cohort study using the TriNetX Research Network, a large federated real-world data platform. We compared outcomes among adults hospitalized with ACS who received NRT alone versus those receiving NRT combined with varenicline or bupropion. Our study aimed to evaluate the cardiovascular safety and effectiveness of combination pharmacotherapy for smoking cessation in the post-ACS population, providing evidence to inform guideline-based care and optimize secondary prevention strategies in routine clinical settings.

## METHODS

### Data source and study design

We performed a retrospective cohort study using the TriNetX Research Network platform (TriNetX, Cambridge, MA), a federated real-world data network that enables analyses across participating healthcare organizations while returning only de-identified results in aggregate form. All cohort building and outcome analyses were conducted within the TriNetX “Compare Outcomes” workflow. Queries were executed on February 16, 2026, using the TriNetX US Collaborative Network, in which 67 healthcare organizations were queried and responded, with contributing providers varying by outcome and data element availability.

### Cohort identification and index event definition

Adults aged 18 years or older were eligible. For all comparisons, we identified patients hospitalized with acute coronary syndrome using inpatient, short-stay, observation, or acute inpatient encounter types and relevant hospitalization services. To support adequate follow-up capture for downstream analyses, the qualifying hospitalization was constrained to occur at least one year before the query execution date within the cohort definition logic. The complete patient selection and matching process is illustrated in **Figure 1**.

**Figure 1:**
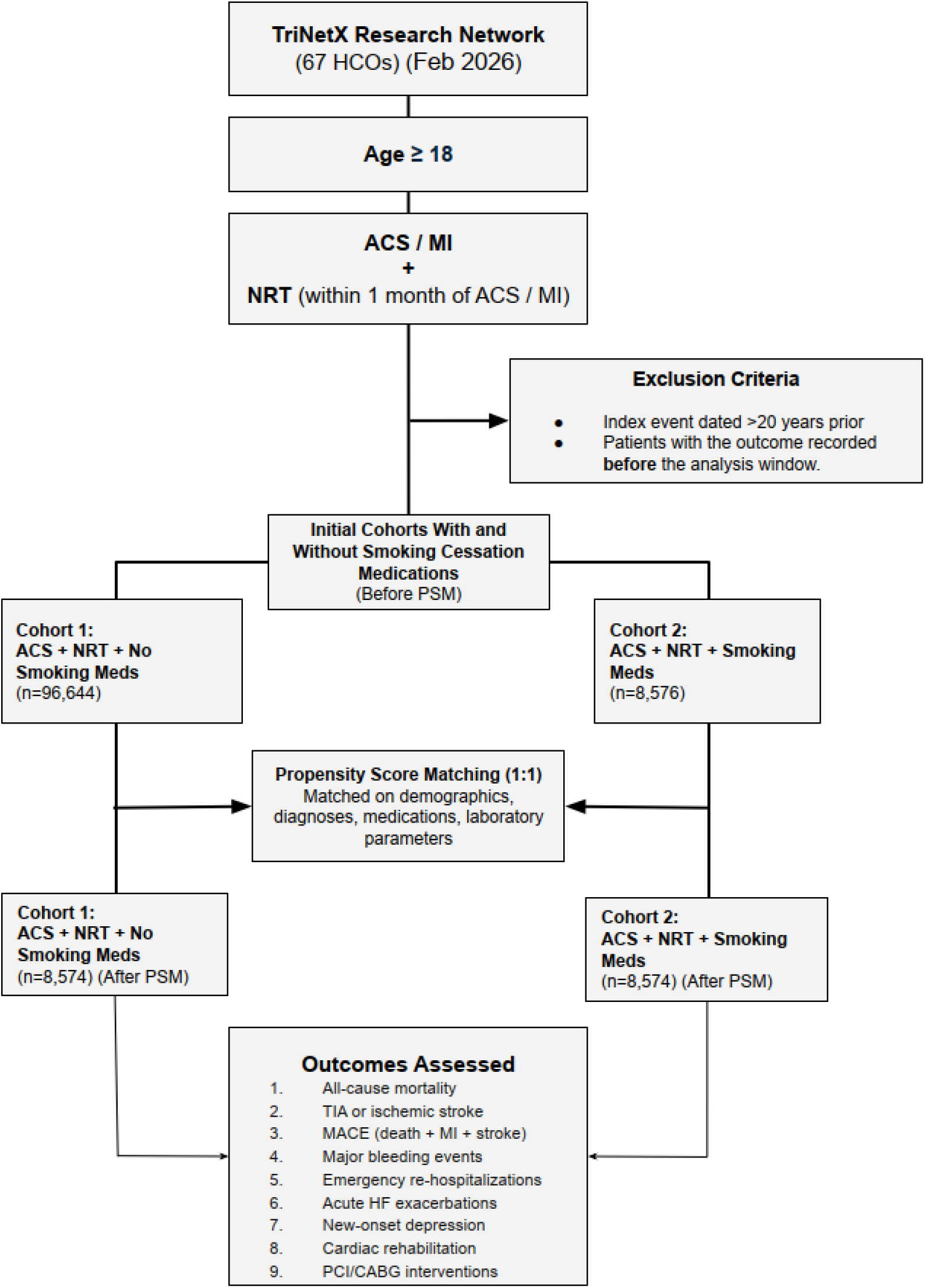
Consort Diagram of Complete Patient Selection Process

### Exposure definitions and comparison groups

The study evaluated prescription smoking cessation pharmacotherapy strategies among patients who received nicotine replacement therapy (NRT) in proximity to the index hospitalization. NRT exposure was defined by either a nicotine replacement procedure code (ICD-10-PCS HZ90) or nicotine medication exposure (RxNorm nicotine). For cohort construction, NRT was required to occur within one month on or after the ACS hospitalization.

We performed three prespecified pairwise comparisons, each anchored on the same ACS hospitalization definition and the same NRT timing requirement. The reference group in all analyses comprised patients with ACS who received NRT within one month of ACS hospitalization but had no evidence of prescription smoking cessation pharmacotherapy, operationalized as the absence of bupropion and varenicline medication exposures. The first comparator group included patients who received bupropion alongside NRT within one month on or after the index hospitalization with no varenicline exposure during that period. The second comparator group included patients who received varenicline alongside NRT within one month on or after the index hospitalization with no bupropion exposure during that period. The third comparator group included patients who received any smoking cessation pharmacotherapy, defined as bupropion or varenicline, alongside NRT within one month on or after the ACS hospitalization. Given that varenicline and bupropion have distinct pharmacologic profiles and may differ in clinical effectiveness, the individual drug comparisons (NRT + varenicline vs. NRT alone and NRT + bupropion vs. NRT alone) were considered co-primary analyses, while the combined pharmacotherapy comparison served as a supportive analysis to assess the overall association of adding any prescription cessation agent to NRT. The index event for each patient was defined using TriNetX’s index event algorithm, in which the index date corresponds to the day on which the patient simultaneously first met all aforementioned cohort-defining criteria. Full eligibility criteria and codes used for cohort construction are provided in **Supplementary Table S1.**

### Follow-up windows

Outcomes were assessed using two follow-up horizons that were analyzed separately. In the 6-month analysis, outcome ascertainment started one day after the index event and extended through 180 days after the index event. In the 1-year analysis, outcome ascertainment started one day after the index event and extended through 365 days after the index event.

### Outcomes

Clinical outcomes were defined using diagnosis, procedure, visit, and medication concepts mapped within TriNetX to standard terminologies including ICD-10-CM, ICD-10-PCS, CPT, HCPCS, and RxNorm. The primary outcome was all-cause mortality. Secondary effectiveness and safety outcomes included transient ischemic attack or ischemic stroke, major bleeding events, emergency rehospitalizations, acute exacerbations of heart failure, a composite major adverse cardiovascular outcome (MACE), cardiac rehabilitation utilization, coronary revascularization procedures (PCI and CABG interventions), and new-onset depression. A falsification endpoint of urinary tract infection was also evaluated as a negative control outcome to assess for residual confounding signals not plausibly related to smoking cessation pharmacotherapy prescription. Detailed outcome definitions and the corresponding codes are provided in **Supplementary Table S2.**

### Covariates and propensity score matching

To reduce confounding, we applied propensity score matching within TriNetX for each comparison using 50 prespecified baseline characteristics spanning demographics, relevant comorbid diagnoses, cardiovascular procedures, baseline medications, and key laboratory or physiologic measurements available in the network. Demographic covariates included age at index, sex, and race categories available in the platform. Clinical covariates included hypertension, type 2 diabetes mellitus, ischemic and hemorrhagic cerebrovascular disease, chronic ischemic heart disease, acute myocardial infarction, atrial fibrillation subtypes, chronic kidney disease, neoplasms, nicotine dependence, alcohol-related disorders, and gastrointestinal hemorrhage. Procedure covariates included coronary interventions captured within the TriNetX procedure ontology. Medication covariates included anticoagulants and antiplatelet therapies commonly used in ACS care, as well as cardiovascular and metabolic medication classes recorded in the platform. Laboratory and physiologic covariates included left ventricular ejection fraction, systolic blood pressure, body mass index, hemoglobin A1c, creatinine, hemoglobin, international normalized ratio (INR), low-density lipoprotein (LDL) cholesterol, and n-terminal pro-B-type natriuretic peptide (NT-proBNP) when available.

Propensity scores were estimated using logistic regression as implemented in TriNetX’s propensity score framework, and matching was conducted in a 1:1 ratio using greedy nearest-neighbor matching without replacement with a default caliper of 0.10 standard deviations, consistent with TriNetX’s documented matching approach. Post-matching balance was assessed using standardized mean differences, with values below 0.1 considered consistent with adequate covariate balance, aligning with common TriNetX reporting conventions.

### Statistical analysis

Baseline characteristics were summarized before and after matching using TriNetX descriptive outputs. For each outcome and follow-up horizon, TriNetX “Measure of Association” outputs were used to estimate absolute risks, risk differences, risk ratios, and odds ratios with 95% confidence intervals for matched cohorts. Time-to-event analyses were conducted using Kaplan-Meier methods within TriNetX, with log-rank testing and hazard ratio estimation where supported by the platform. In survival analyses, censoring followed the TriNetX approach in which individuals are censored after the last recorded encounter during the study period. All statistical tests were two-sided, and statistical significance was assessed at an alpha level of 0.05. Given the number of outcomes examined, all-cause mortality was designated as the single primary endpoint, and all remaining outcomes were considered secondary or exploratory. No formal correction for multiplicity of statistical testing was applied; however, results for secondary outcomes should be interpreted in the context of multiple comparisons, and emphasis is placed on the consistency of effect direction and magnitude rather than individual p-values. This study was conducted and reported in accordance with the Strengthening the Reporting of Observational Studies in Epidemiology (STROBE) guidelines.

### Ethics and data privacy

TriNetX provides de-identified data in accordance with the HIPAA Privacy Rule de-identification standard, attested via qualified expert determination, and maintains an information security management system aligned with ISO 27001:2022 as described in TriNetX publication guidance. Because this study used de-identified, aggregate-level data with no access to direct identifiers, it is considered not human subjects research under U.S. regulatory definitions, and the UTMB IRB confirmed use of this data as exempt from IRB review.

## RESULTS

### Study Sample and Propensity Score Matching

For the 1-year follow-up analyses, the NRT-only comparator cohort comprised 97,084 patients prior to matching; the corresponding prescription pharmacotherapy cohorts included 8,629 patients. After 1:1 nearest-neighbor propensity score matching on 50 covariates with a caliper of 0.10 standard deviations, the matched cohorts comprised 4,654 pairs in the bupropion analysis, 2,126 pairs in the varenicline analysis, and 8,574 pairs in the combined analysis. Before matching, several clinically meaningful imbalances were present: the pharmacotherapy group was younger (mean age 57.3 ± 10.4 vs. 58.7 ± 11.8 years; standardized mean difference [SMD] = 0.124), more likely to be female (43.0% vs. 35.9%; SMD = 0.145), and had higher prevalences of hypertensive diseases (75.9% vs. 60.6%; SMD = 0.332), type 2 diabetes mellitus (37.0% vs. 27.2%; SMD = 0.211), chronic ischemic heart disease (71.7% vs. 50.0%; SMD = 0.458), and acute myocardial infarction (74.6% vs. 59.1%; SMD = 0.333). After matching, all standardized mean differences fell below 0.10, confirming adequate covariate balance **(Table 1)**. Mean age was 57.3 years in both cohorts (SMD = 0.002), and the proportion of female patients was 42.3% versus 43.0% (SMD = 0.013). Race and ethnicity distributions were comparable, with White patients comprising 80.9% and 81.3% of the NRT-only and pharmacotherapy groups, respectively (SMD = 0.009), and Black or African American patients comprising 13.1% in both cohorts (SMD = 0.002). Key cardiovascular comorbidities were well balanced, including hypertensive diseases (75.0% vs. 75.9%; SMD = 0.020), type 2 diabetes mellitus (36.8% vs. 37.0%; SMD = 0.003), chronic ischemic heart disease (71.8% vs. 71.7%; SMD = 0.002), and acute myocardial infarction (74.8% vs. 74.6%; SMD = 0.004). Baseline cardiovascular medication use was similarly distributed: aspirin (72.5% vs. 72.6%; SMD = 0.001), clopidogrel (35.6% vs. 34.9%; SMD = 0.015), ticagrelor (10.5% vs. 10.0%; SMD = 0.014), and prasugrel (4.4% in both; SMD < 0.001). Complete pre- and post-matching covariate distributions are presented in **Supplementary Table S3**.

**Table 1:**
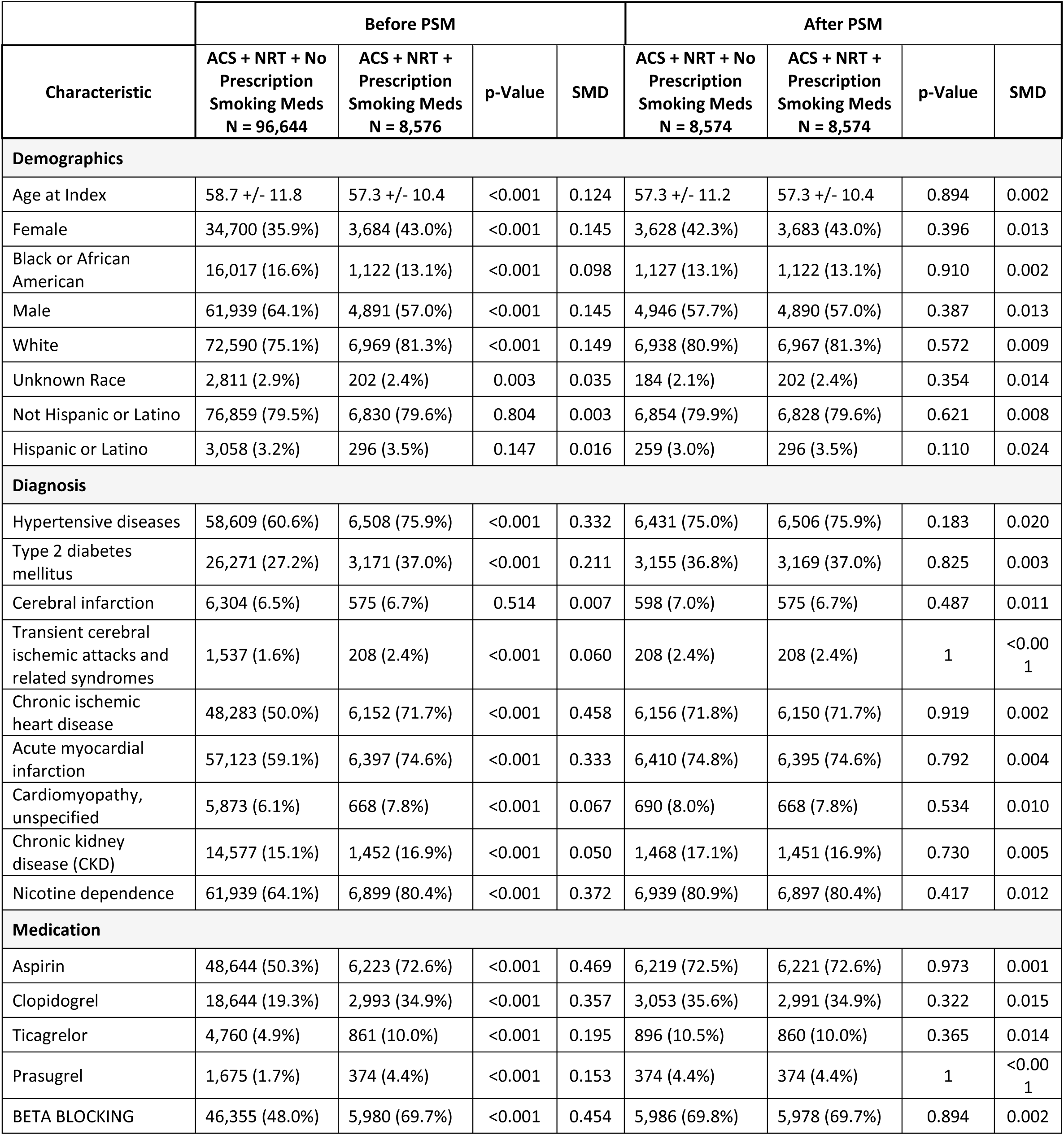
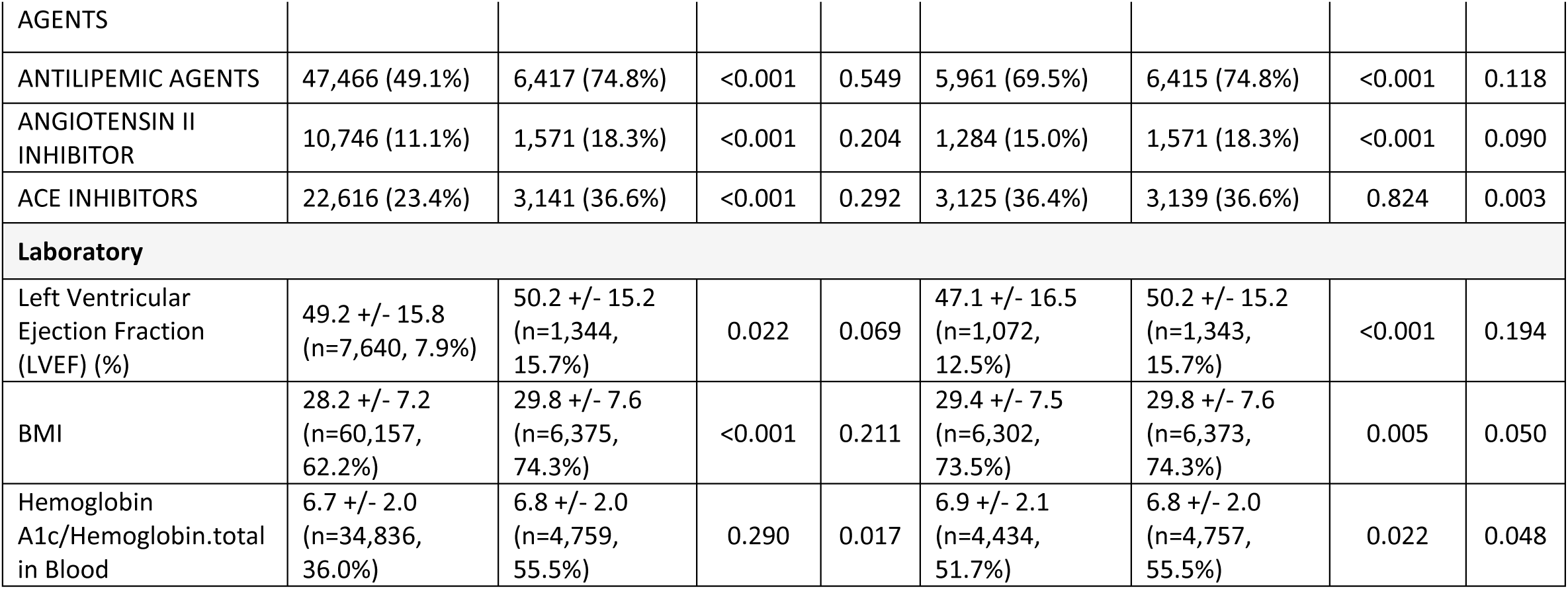
Selected baseline characteristics of the study cohorts before and after propensity score matching.

Throughout the following results, hazard ratios (HRs) greater than 1.0 indicate a higher hazard in the NRT-only group relative to the pharmacotherapy group, while HRs less than 1.0 indicate a higher hazard in the pharmacotherapy group.

### Co-Primary Analysis 1: Prescription Bupropion + NRT vs. NRT Alone

At 6 months, the NRT-only group had a significantly higher hazard of all-cause mortality compared with the bupropion group (HR 1.171, 95% CI 1.039-1.320, p = 0.009). Significant differences favoring the bupropion group were also observed for major bleeding events (HR 1.175, 95% CI 1.050-1.314, p = 0.005) and MACE (HR 1.135, 95% CI 1.076-1.198, p < 0.001). Cardiac rehabilitation utilization was significantly greater in the bupropion group (HR 0.798, 95% CI 0.686-0.928, p = 0.003). However, the bupropion group had significantly more emergency rehospitalizations (HR 0.906, 95% CI 0.854-0.963, p = 0.001). No significant differences were observed for TIA or ischemic stroke (HR 0.883, 95% CI 0.759-1.026, p = 0.103) or acute HF exacerbations (HR 1.100, 95% CI 0.993-1.219, p = 0.068) **(Table 2)**.

**Table 2:**
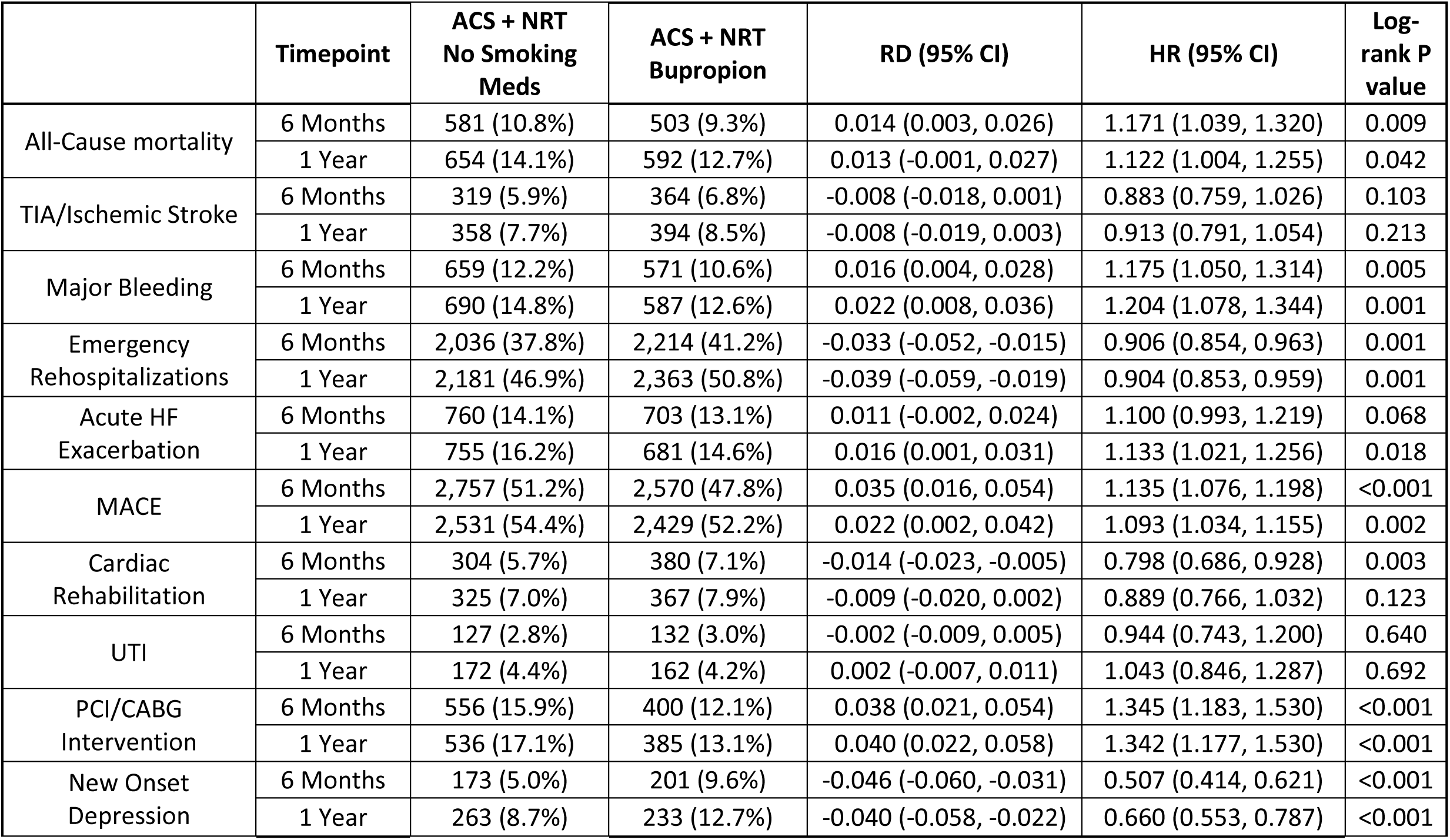
Comparison of clinical outcomes after propensity score matching: Bupropion + NRT versus NRT Alone (Co-Primary Analysis 1)

|  | Timepoint | ACS + NRT<br>No Smoking<br>Meds | ACS + NRT<br>Bupropion | RD (95% CI) | HR (95% CI) | Log-rank P<br>value |
| --- | --- | --- | --- | --- | --- | --- |
| All-Cause mortality | 6 Months | 581 (10.8%) | 503 (9.3%) | 0.014 (0.003, 0.026) | 1.171 (1.039, 1.320) | 0.009 |
|  | 1 Year | 654 (14.1%) | 592 (12.7%) | 0.013 (-0.001, 0.027) | 1.122 (1.004, 1.255) | 0.042 |
| TIA/Ischemic Stroke | 6 Months | 319 (5.9%) | 364 (6.8%) | -0.008 (-0.018, 0.001) | 0.883 (0.759, 1.026) | 0.103 |
|  | 1 Year | 358 (7.7%) | 394 (8.5%) | -0.008 (-0.019, 0.003) | 0.913 (0.791, 1.054) | 0.213 |
| Major Bleeding | 6 Months | 659 (12.2%) | 571 (10.6%) | 0.016 (0.004, 0.028) | 1.175 (1.050, 1.314) | 0.005 |
|  | 1 Year | 690 (14.8%) | 587 (12.6%) | 0.022 (0.008, 0.036) | 1.204 (1.078, 1.344) | 0.001 |
| Emergency<br>Rehospitalizations | 6 Months | 2,036 (37.8%) | 2,214 (41.2%) | -0.033 (-0.052, -0.015) | 0.906 (0.854, 0.963) | 0.001 |
|  | 1 Year | 2,181 (46.9%) | 2,363 (50.8%) | -0.039 (-0.059, -0.019) | 0.904 (0.853, 0.959) | 0.001 |
| Acute HF<br>Exacerbation | 6 Months | 760 (14.1%) | 703 (13.1%) | 0.011 (-0.002, 0.024) | 1.100 (0.993, 1.219) | 0.068 |
|  | 1 Year | 755 (16.2%) | 681 (14.6%) | 0.016 (0.001, 0.031) | 1.133 (1.021, 1.256) | 0.018 |
| MACE | 6 Months | 2,757 (51.2%) | 2,570 (47.8%) | 0.035 (0.016, 0.054) | 1.135 (1.076, 1.198) | <0.001 |
|  | 1 Year | 2,531 (54.4%) | 2,429 (52.2%) | 0.022 (0.002, 0.042) | 1.093 (1.034, 1.155) | 0.002 |
| Cardiac<br>Rehabilitation | 6 Months | 304 (5.7%) | 380 (7.1%) | -0.014 (-0.023, -0.005) | 0.798 (0.686, 0.928) | 0.003 |
|  | 1 Year | 325 (7.0%) | 367 (7.9%) | -0.009 (-0.020, 0.002) | 0.889 (0.766, 1.032) | 0.123 |
| UTI | 6 Months | 127 (2.8%) | 132 (3.0%) | -0.002 (-0.009, 0.005) | 0.944 (0.743, 1.200) | 0.640 |
|  | 1 Year | 172 (4.4%) | 162 (4.2%) | 0.002 (-0.007, 0.011) | 1.043 (0.846, 1.287) | 0.692 |
| PCI/CABG<br>Intervention | 6 Months | 556 (15.9%) | 400 (12.1%) | 0.038 (0.021, 0.054) | 1.345 (1.183, 1.530) | <0.001 |
|  | 1 Year | 536 (17.1%) | 385 (13.1%) | 0.040 (0.022, 0.058) | 1.342 (1.177, 1.530) | <0.001 |
| New Onset<br>Depression | 6 Months | 173 (5.0%) | 201 (9.6%) | -0.046 (-0.060, -0.031) | 0.507 (0.414, 0.621) | <0.001 |
|  | 1 Year | 263 (8.7%) | 233 (12.7%) | -0.040 (-0.058, -0.022) | 0.660 (0.553, 0.787) | <0.001 |

At 1 year, the mortality benefit persisted (HR 1.122, 95% CI 1.004-1.255, p = 0.042). The NRT-only group continued to have a higher hazard of major bleeding (HR 1.204, 95% CI 1.078-1.344, p = 0.001) and MACE (HR 1.093, 95% CI 1.034-1.155, p = 0.002). Acute HF exacerbations became significant at 1 year, with a higher hazard in the NRT-only group (HR 1.133, 95% CI 1.021-1.256, p = 0.018). The bupropion group continued to have significantly more emergency rehospitalizations (HR 0.904, 95% CI 0.853-0.959, p = 0.001). No significant differences were found for TIA or ischemic stroke (HR 0.913, 95% CI 0.791-1.054, p = 0.213) or cardiac rehabilitation utilization (HR 0.889, 95% CI 0.766-1.032, p = 0.123) **(Figure 2)**.

**Figure 2:**
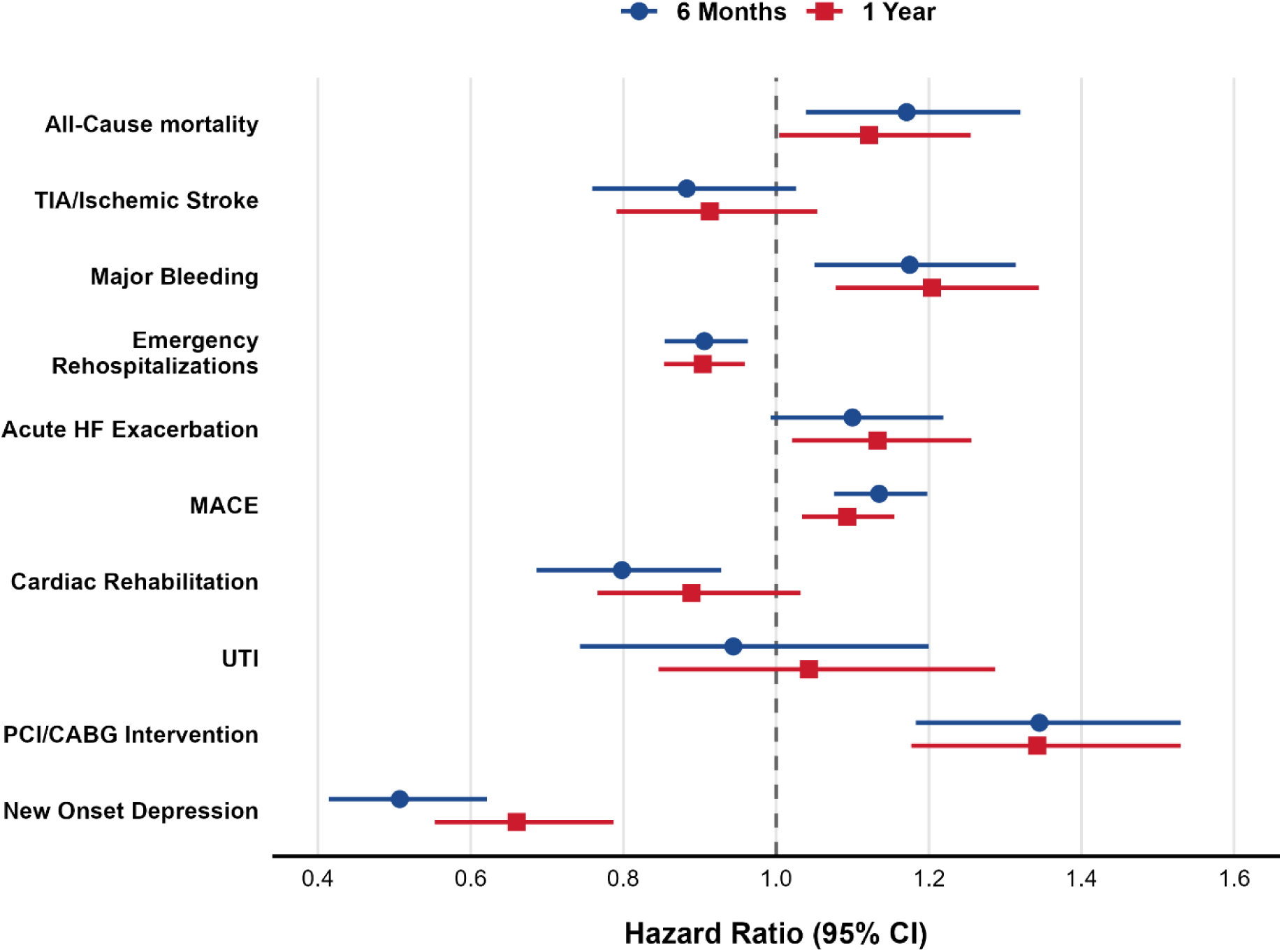
Forest Plot of Hazard Ratios for All Clinical Outcomes After Propensity Score Matching: Bupropion + NRT versus NRT Alone (Co-Primary Analysis 1)

### Co-Primary Analysis 2: Prescription Varenicline + NRT vs. NRT Alone

At 6 months, the NRT-only group had a significantly higher hazard of all-cause mortality (HR 1.301, 95% CI 1.042-1.626, p = 0.020), major bleeding events (HR 1.348, 95% CI 1.095-1.661, p = 0.005), acute HF exacerbations (HR 1.489, 95% CI 1.257-1.765, p < 0.001), and MACE (HR 1.233, 95% CI 1.132-1.343, p < 0.001). Emergency rehospitalizations were also significantly higher in the NRT-only group (HR 1.116, 95% CI 1.009-1.234, p = 0.033). No significant differences were observed for TIA or ischemic stroke (HR 1.171, 95% CI 0.915-1.499, p = 0.208) or cardiac rehabilitation utilization (HR 0.838, 95% CI 0.685-1.026, p = 0.087) **(Table 3)**. At 1 year, the mortality benefit persisted (HR 1.228, 95% CI 1.017-1.483, p = 0.032). Significant differences favoring the varenicline group continued for major bleeding (HR 1.223, 95% CI 1.018-1.469, p = 0.031), HF exacerbations (HR 1.418, 95% CI 1.212-1.659, p < 0.001), and MACE (HR 1.187, 95% CI 1.094-1.288, p < 0.001). Emergency rehospitalizations were no longer significantly different at 1 year (HR 1.036, 95% CI 0.948-1.133, p = 0.435). TIA or ischemic stroke (HR 1.160, 95% CI 0.930-1.446, p = 0.188) and cardiac rehabilitation utilization (HR 0.825, 95% CI 0.677-1.006, p = 0.056) remained non-significant **(Figure 3)**.

**Figure 3:**
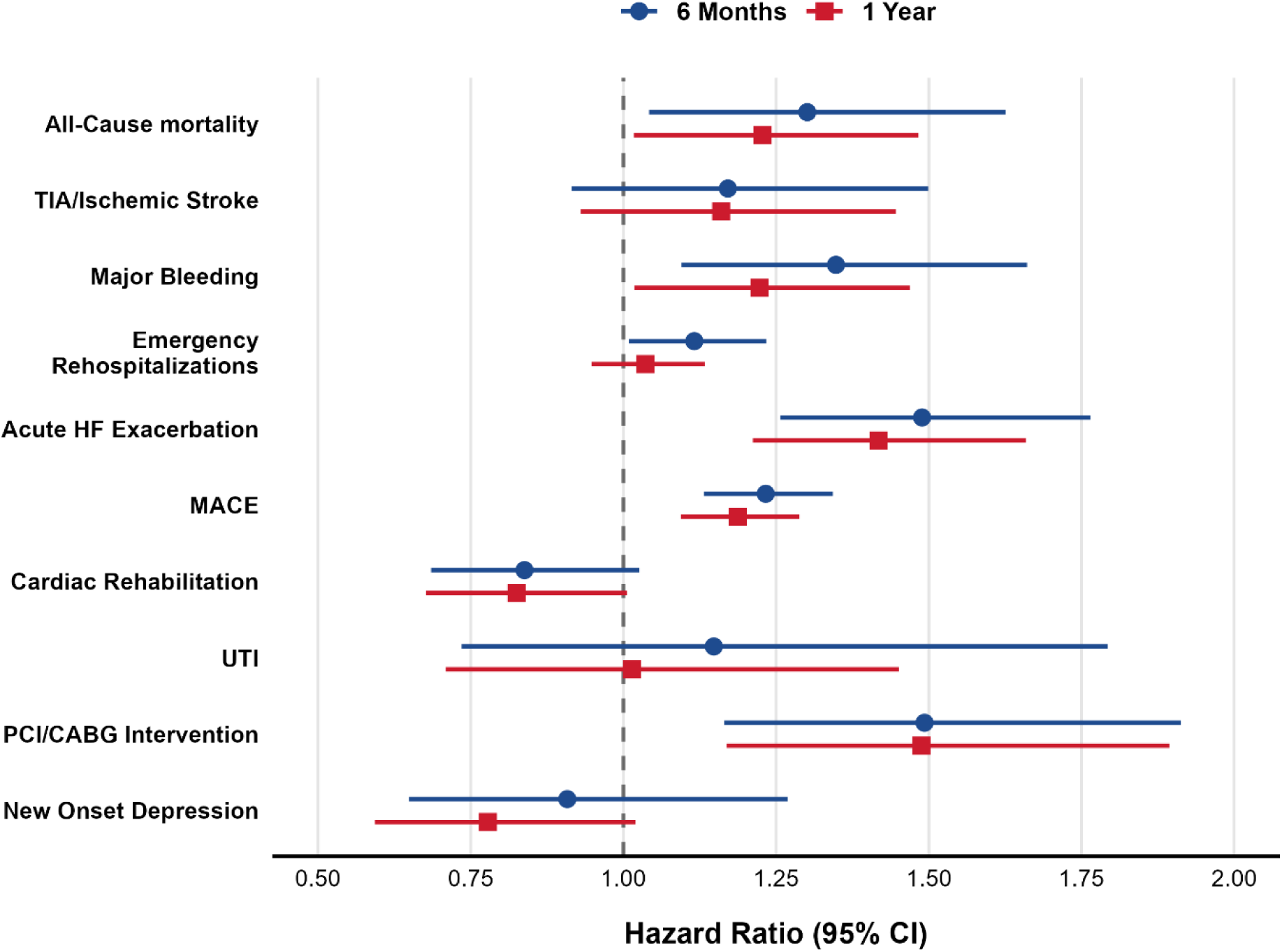
Forest Plot of Hazard Ratios for All Clinical Outcomes After Propensity Score Matching: Varenicline + NRT versus NRT Alone (Co-Primary Analysis 2)

**Table 3:** Comparison of clinical outcomes after propensity score matching: Varenicline + NRT versus NRT Alone (Co-Primary Analysis 2)

|  | Timepoint | ACS + NRT<br>No Smoking<br>Meds | ACS + NRT<br>Varenicline | RD (95% CI) | HR (95% CI) | Log-rank P<br>value |
| --- | --- | --- | --- | --- | --- | --- |
| All-Cause mortality | 6 Months | 175 (8.2%) | 139 (6.5%) | 0.017 (0.001, 0.033) | 1.301 (1.042, 1.626) | 0.020 |
|  | 1 Year | 236 (11.1%) | 199 (9.4%) | 0.017 (-0.001, 0.036) | 1.228 (1.017, 1.483) | 0.032 |
| TIA/Ischemic Stroke | 6 Months | 135 (6.3%) | 119 (5.6%) | 0.008 (-0.007, 0.022) | 1.171 (0.915, 1.499) | 0.208 |
|  | 1 Year | 167 (7.9%) | 149 (7.0%) | 0.008 (-0.007, 0.024) | 1.160 (0.930, 1.446) | 0.188 |
| Major Bleeding | 6 Months | 203 (9.5%) | 157 (7.4%) | 0.022 (0.005, 0.038) | 1.348 (1.095, 1.661) | 0.005 |
|  | 1 Year | 248 (11.7%) | 212 (10.0%) | 0.017 (-0.002, 0.036) | 1.223 (1.018, 1.469) | 0.031 |
| Emergency<br>Rehospitalizations | 6 Months | 774 (36.4%) | 737 (34.7%) | 0.017 (-0.011, 0.046) | 1.116 (1.009, 1.234) | 0.033 |
|  | 1 Year | 946 (44.5%) | 974 (45.8%) | -0.013 (-0.043, 0.017) | 1.036 (0.948, 1.133) | 0.435 |
| Acute HF<br>Exacerbation | 6 Months | 322 (15.1%) | 228 (10.7%) | 0.044 (0.024, 0.064) | 1.489 (1.257, 1.765) | <0.001 |
|  | 1 Year | 364 (17.1%) | 272 (12.8%) | 0.043 (0.022, 0.065) | 1.418 (1.212, 1.659) | <0.001 |
| MACE | 6 Months | 1,105 (52.0%) | 999 (47.0%) | 0.050 (0.020, 0.080) | 1.233 (1.132, 1.343) | <0.001 |
|  | 1 Year | 1,198 (56.3%) | 1,125 (52.9%) | 0.034 (0.004, 0.064) | 1.187 (1.094, 1.288) | <0.001 |
| Cardiac<br>Rehabilitation | 6 Months | 171 (8.0%) | 208 (9.8%) | -0.017 (-0.035, -0.000) | 0.838 (0.685, 1.026) | 0.087 |
|  | 1 Year | 178 (8.4%) | 220 (10.3%) | -0.020 (-0.037, -0.002) | 0.825 (0.677, 1.006) | 0.056 |
| UTI | 6 Months | 41 (2.2%) | 35 (2.0%) | 0.003 (-0.006, 0.012) | 1.148 (0.735, 1.793) | 0.545 |
|  | 1 Year | 59 (3.2%) | 57 (3.2%) | 0.000 (-0.011, 0.012) | 1.014 (0.709, 1.451) | 0.939 |
| PCI/CABG<br>Intervention | 6 Months | 159 (14.1%) | 103 (9.9%) | 0.041 (0.014, 0.069) | 1.493 (1.165, 1.913) | 0.001 |
|  | 1 Year | 167 (14.8%) | 109 (10.5%) | 0.043 (0.015, 0.071) | 1.488 (1.169, 1.894) | 0.001 |
| New Onset<br>Depression | 6 Months | 66 (4.6%) | 71 (5.3%) | -0.007 (-0.023, 0.009) | 0.908 (0.649, 1.269) | 0.572 |
|  | 1 Year | 94 (6.6%) | 118 (8.8%) | -0.022 (-0.042, -0.002) | 0.778 (0.593, 1.020) | 0.068 |

### Supportive Analysis: Combined Pharmacotherapy + NRT vs. NRT Alone

At 6 months, the NRT-only group had a significantly higher hazard of all-cause mortality (HR 1.240, 95% CI 1.125-1.366, p < 0.001), major bleeding (HR 1.224, 95% CI 1.116-1.342, p < 0.001), HF exacerbations (HR 1.158, 95% CI 1.067-1.256, p < 0.001), and MACE (HR 1.157, 95% CI 1.109-1.206, p < 0.001). Cardiac rehabilitation utilization was significantly greater in the pharmacotherapy group (HR 0.847, 95% CI 0.761-0.944, p = 0.003). No significant differences were found for TIA or ischemic stroke (HR 0.998, 95% CI 0.890-1.120, p = 0.975) or emergency rehospitalizations (HR 0.987, 95% CI 0.941-1.034, p = 0.574) **(Supplementary Table S4)**.

At 1 year, results were consistent. All-cause mortality (HR 1.257, 95% CI 1.155-1.369, p < 0.001), major bleeding (HR 1.180, 95% CI 1.084-1.284, p < 0.001), HF exacerbations (HR 1.160, 95% CI 1.075-1.252, p < 0.001), MACE (HR 1.164, 95% CI 1.118-1.213, p < 0.001), and cardiac rehabilitation utilization (HR 0.824, 95% CI 0.740-0.916, p < 0.001) all favored the pharmacotherapy group. TIA or ischemic stroke approached but did not reach significance (HR 1.104, 95% CI 0.995-1.225, p = 0.062), and emergency rehospitalizations showed no significant difference (HR 0.969, 95% CI 0.928-1.011, p = 0.146).

### Sensitivity Analyses

In the PCI/CABG sensitivity analysis, which excluded patients with prior revascularization procedures, the NRT-only group had a significantly higher hazard of undergoing new PCI or CABG across all comparisons at both time points. At 1 year, HRs were 1.342 (95% CI 1.177-1.530, p < 0.001) for bupropion, 1.488 (95% CI 1.169-1.894, p = 0.001) for varenicline, and 1.393 (95% CI 1.257-1.544, p < 0.001) for combined pharmacotherapy. At 6 months, findings were consistent: HR 1.345 (95% CI 1.183-1.530, p < 0.001) for bupropion, HR 1.493 (95% CI 1.165-1.913, p = 0.001) for varenicline, and HR 1.373 (95% CI 1.236-1.525, p < 0.001) for the combined analysis **(Tables 2, 3, and Supplementary Table S4)**.

In the new-onset depression sensitivity analysis, which excluded patients with prior depression diagnoses, the pharmacotherapy group had a significantly higher hazard of new depression diagnoses in the bupropion and combined comparisons. At 1 year, HRs were 0.660 (95% CI 0.553-0.787, p < 0.001) for bupropion, 0.778 (95% CI 0.593-1.020, p = 0.068) for varenicline, and 0.676 (95% CI 0.592-0.771, p < 0.001) for the combined analysis. At 6 months, this pattern was more pronounced for bupropion (HR 0.507, 95% CI 0.414-0.621, p < 0.001) and the combined group (HR 0.693, 95% CI 0.596-0.806, p < 0.001), whereas the varenicline comparison was not significant (HR 0.908, 95% CI 0.649-1.269, p = 0.572) **(Tables 2, 3, and Supplementary Table S4)**.

### Falsification Endpoint

Urinary tract infection, assessed as a negative control outcome, showed no significant differences between groups in any comparison at either time point (all p > 0.40), supporting the validity of the propensity score matching **(Tables 2, 3, and Supplementary Table S4)**. At 1 year, risk ratios were 1.043 (95% CI 0.846-1.287, p = 0.692) for bupropion, 1.014 (95% CI 0.709-1.451, p = 0.939) for varenicline, and 1.050 (95% CI 0.897-1.229, p = 0.547) for the combined analysis.

## DISCUSSION

In this large real-world analysis of patients hospitalized with ACS who initiated nicotine replacement therapy (NRT), the addition of smoking cessation pharmacotherapy, either varenicline or bupropion, was associated with significantly lower mortality, major bleeding, heart failure exacerbations, and major adverse cardiovascular events (MACE) compared with NRT alone over both 6-month and 1-year follow-up. Importantly, these benefits were not associated with an increased risk of cerebrovascular events and were accompanied by greater engagement in cardiac rehabilitation. Although smoking abstinence could not be directly verified in this dataset, these findings are most plausibly mediated by higher rates of successful smoking cessation among patients receiving pharmacotherapy, consistent with prior evidence demonstrating superior quit rates with varenicline and bupropion compared with NRT alone. Together, these findings suggest that pharmacotherapy-assisted smoking cessation strategies may confer clinically meaningful cardiovascular benefits beyond those achieved with NRT alone in high-risk patients following ACS.

### Smoking cessation and cardiovascular risk after ACS

Cigarette smoking remains a major contributor to atherosclerotic cardiovascular disease and is responsible for substantial excess mortality among patients with established coronary artery disease (1,2). Continued smoking after ACS is strongly associated with recurrent ischemic events, progression of heart failure, and premature death (4,5). Observational studies suggest that smoking cessation after myocardial infarction reduces mortality by approximately one-third, making it one of the most effective interventions in secondary prevention (6,13). Accordingly, contemporary cardiovascular prevention guidelines emphasize smoking cessation following ACS as a Class I recommendation (14,15). Despite these recommendations, relapse rates remain substantial, particularly when cessation efforts rely solely on behavioral counseling or NRT monotherapy(16,17).

The early post-ACS period represents a meaningful opportunity for smoking cessation interventions. Patients hospitalized for ACS often demonstrate increased motivation to quit, and structured cessation programs initiated during hospitalization have been associated with improved abstinence rates and long-term cardiovascular outcomes (7,18). However, despite this window of opportunity, the implementation of pharmacologic smoking cessation therapy remains suboptimal in routine clinical practice, partly due to persistent concerns regarding cardiovascular safety in patients with recent acute events (3,5).

### Comparative effectiveness of smoking cessation pharmacotherapy

In the general smoking population, randomized trials have demonstrated superior abstinence rates with varenicline or bupropion compared with NRT alone (19,20). Varenicline, a partial agonist at the α4β2 nicotinic acetylcholine receptor, reduces nicotine cravings and withdrawal while simultaneously attenuating the reinforcing effects of nicotine (19). Bupropion, a norepinephrine-dopamine reuptake inhibitor with nicotinic receptor antagonist properties, enhances dopaminergic reward pathways while reducing withdrawal symptoms (20). Both agents have demonstrated efficacy in large randomized trials and meta-analyses evaluating smoking cessation interventions (19–22).

However, the translation of these findings into populations with recent cardiovascular events has been slower. Early post-marketing reports raised concerns about potential cardiovascular and neuropsychiatric adverse effects of these medications, particularly varenicline, which resulted in caution among clinicians treating patients with recent ACS (23). Earlier safety concerns regarding varenicline and bupropion have been largely refuted by subsequent evidence. Large randomized trials, including the EAGLES trial, along with observational studies, have demonstrated no significant increase in cardiovascular or neuropsychiatric adverse events, providing strong reassurance regarding their safety in patients with cardiovascular disease (24–26).

Our findings extend this evidence base by demonstrating that pharmacotherapy-assisted smoking cessation strategies, associated with presumed higher rates of smoking cessation, are linked to favorable clinical cardiovascular outcomes in real-world practice. Patients receiving varenicline or bupropion in addition to NRT experienced lower mortality and fewer cardiovascular events compared with those receiving NRT alone, suggesting that more intensive cessation strategies may translate into measurable improvements in clinical outcomes.

### Cardiovascular safety of varenicline and bupropion

The cardiovascular safety of smoking cessation pharmacotherapies has been evaluated in several prior studies. The Evaluating Adverse Events in a Global Smoking Cessation Study (EAGLES) and subsequent randomized investigations found no significant increase in major cardiovascular events among patients treated with varenicline, bupropion, or nicotine patch compared with placebo (24). Similarly, large meta-analyses have shown that varenicline does not increase the risk of cardiovascular events in smokers with or without underlying cardiovascular disease (25,26).

Our analysis provides complementary real-world evidence supporting the cardiovascular safety of these therapies in the context of recent ACS and suggests that combining NRT with prescription pharmacotherapy does not confer additional cardiovascular risk, addressing a relative gap in the existing literature. Notably, neither varenicline nor bupropion was associated with increased rates of transient ischemic attack or ischemic stroke in our cohorts, and both agents were associated with lower rates of major bleeding events compared with NRT alone. The observed reduction in bleeding risk may reflect indirect effects mediated by smoking cessation itself, as continued smoking is associated with increased platelet activation, endothelial dysfunction, and inflammatory signaling that contribute to vascular instability (27,28). Smoking cessation has been shown to improve endothelial function, reduce platelet aggregation, and normalize prothrombotic pathways within weeks of quitting (29).

### Potential mechanisms underlying improved outcomes

Several mechanisms may explain the favorable outcomes observed among patients receiving smoking cessation pharmacotherapy in our study. First, pharmacotherapy is known to significantly increase the likelihood of sustained smoking abstinence compared with NRT alone (21,22). Sustained cessation reduces sympathetic activation, oxidative stress, and inflammatory signaling, all of which contribute to atherosclerotic plaque instability and recurrent ischemic events (28–30). Smoking cessation also leads to improvements in endothelial function and reductions in thrombogenicity, which may explain the lower rates of MACE observed in pharmacotherapy recipients (29,30).

Second, pharmacotherapy-assisted smoking cessation was associated with higher utilization of cardiac rehabilitation. While the directionality of this association cannot be determined, it is possible that patients who are more engaged in their recovery are both more likely to attend rehabilitation and to use cessation medications, or that rehabilitation enhances understanding and effective use of these therapies. Regardless, participation in cardiac rehabilitation is independently associated with lower mortality, improved functional capacity, and reduced cardiovascular events after ACS, suggesting that increased engagement with rehabilitation may be one pathway through which pharmacotherapy contributes to improved long-term outcomes (31,32).

Third, nicotine exposure itself may influence cardiovascular physiology. Although NRT is generally considered safe, nicotine has sympathomimetic effects that may increase heart rate, blood pressure, and myocardial oxygen demand (33,34).

### Differences between varenicline and bupropion

Subgroup analyses suggested broadly consistent benefits across pharmacotherapies, though varenicline, generally more effective for smoking cessation, was associated with lower mortality and bleeding at 6 months and sustained reductions in MACE and heart failure at 1 year. Bupropion is often prescribed for depression, so some patients may not have used it for cessation. Similar effects observed between agents may reflect residual confounding rather than true equivalence. Bupropion was associated with reductions in bleeding and MACE and greater cardiac rehabilitation utilization, which may reflect both its effects on smoking cessation and potential benefits related to underlying mental health, as it is commonly prescribed for depression. These findings align with prior literature suggesting that varenicline may produce higher smoking abstinence rates compared with other pharmacologic strategies, while bupropion provides a useful alternative when varenicline is contraindicated or not tolerated (21,35).

### Clinical implications

These findings have important implications for clinical practice. Despite strong guideline recommendations for smoking cessation after ACS, pharmacologic treatment remains underutilized in many healthcare settings (3,36). Our findings suggest that clinicians should consider early initiation of combination smoking cessation pharmacotherapy in addition to NRT when appropriate, as this approach may improve not only abstinence rates but also clinically meaningful cardiovascular outcomes.

From a systems perspective, hospitalization for ACS may represent an ideal opportunity to initiate structured smoking cessation programs that incorporate pharmacotherapy, behavioral counseling, and follow-up support (37). Integrating smoking cessation pharmacotherapy within the post-ACS care pathways and discharge protocols, may therefore represent a scalable strategy to improve secondary prevention outcomes.

### Limitations

The primary limitation of this study is that smoking status and abstinence outcomes could not be directly verified in the electronic health records, meaning the observed association between pharmacotherapy use and clinical outcomes is inferred rather than directly confirmed. Differences in quit rates between groups could not be quantified, and important behavioral factors such as motivation to quit, intensity of counseling, and adherence to cessation strategies were unmeasured, all of which may have influenced both treatment selection and outcomes. Additional limitations include incomplete capture of care delivered outside participating healthcare systems, particularly outpatient services like cardiac rehabilitation; residual confounding despite extensive propensity score matching across 50 covariates; reliance on prescription records without confirmation of actual medication use; heterogeneity due to variable coding and follow-up across sites; and restriction of outcomes to the first year post-ACS, leaving longer-term effects uncertain.

## Conclusion

In this large propensity-matched real-world analysis of patients hospitalized with ACS, the addition of smoking cessation pharmacotherapy such as bupropion or varenicline, to nicotine replacement therapy was associated with lower mortality, reduced bleeding events, fewer heart failure exacerbations, and lower rates of major adverse cardiovascular events compared with NRT alone. These findings support the cardiovascular safety of varenicline and bupropion in the post-ACS setting and suggest that more intensive pharmacologic smoking cessation strategies may confer clinically meaningful benefits. Broader integration of pharmacotherapy-assisted smoking cessation into post-ACS care pathways may therefore represent an important opportunity to improve secondary cardiovascular prevention.

## Data Availability

All data supporting the findings of this study are included within the manuscript and its supplementary materials

## Acknowledgment

We are grateful to the TriNetX platform for providing the database and the analysis tools. For additional information, please refer to https://trinetx.com/.

## Ethics Consideration

Due to the de-identified nature of the platform’s patient data and HCOs in compliance with Section §164.514 of the HIPAA Privacy Rule, ethical approval was deemed unnecessary. The study complied with the ethical and reporting standards depicted by the TriNetX platform under “Publication Guidelines”.

## Consent to Participate

Not applicable.

## Consent for publication

Not applicable.

## Availability of data and materials

The authors are not allowed to share the database. Further inquiries should be directed to the corresponding author.

## Animal Ethics

All authors have confirmed that this study did not involve animal subjects or tissue.

